# Factors associated with limited access to condoms and sources of condoms during the COVID-19 pandemic in South Africa

**DOI:** 10.1101/2020.09.11.20192849

**Authors:** Obasanjo Afolabi Bolarinwa

## Abstract

**Background:** Evidence has shown that the prescribed lockdown and physical distancing due to the novel coronavirus disease 2019 (COVID-19) have made accessing essential health care services much more difficult in low-and middle-income countries. Access to contraception is an essential service and should not be denied, even in a global crisis, because of its associated health benefits. Therefore, it is important to maintain timely access to contraception without unnecessary barriers. Hence, this study examines the factors contributing to limited access to condoms and preferred source of condoms during the COVID-19 pandemic in South Africa.

**Methods:** This study used data from the National Income Dynamics Study-Coronavirus Rapid Mobile Survey (NIDS-CRAM) wave 1 survey. The NIDS-CRAM is a nationally representative survey of the National Income Dynamics Survey (NIDS), which involves a sample of South Africans from 2017 NIDS wave 5, who were then re□interviewed via telephone interview. This is the first secondary dataset on coronavirus from NIDS during the coronavirus pandemic. A total of 5,304 respondents were included in the study. Data were analysed using frequencies and percentages, chi-square test and binary logistic regression analysis.

**Results:** Almost one-quarter (22.40%) of South Africans could not access condoms, and every 7 in 10 South Africans preferred public source of condoms. Those who were other population groups [aOR=0.37; 95% CI=0.19-0.74] and those who were in the third wealth quintile [aOR=0.60; 95% CI=0.38-0.93] had lower odds of having access to condoms while those respondents who were aged 25-34 [aOR=0.48; 95% CI=0.27-0.83] and those with a secondary level of education and above [aOR=0.24; 95% CI=0.08-0.71] were less likely to prefer public source of condom.

**Conclusion:** This study concludes that there was limited access to condoms during the COVID-19 pandemic and that the preferred source of condoms was very skewed to public source in South Africa. Strategic interventions such as community distribution of free condoms to avert obstruction of condom access during the COVID-19 pandemic or any future pandemics should be adopted.

## Background

The highly contagious coronavirus disease 2019 (COVID-19) outbreak has revealed how strikingly unprepared the world is for a pandemic and how easily viruses spread in our interconnected world, which has radically changed social relations in the world (1-3). The first cases of SARS-CoV-2 were declared in Africa in late February and early March 2020 (4). South Africa (SA) had its first case reported on March 06, 2020 (5); since then, cases have increased to over 1,170,590 and more than 31,809 deaths have been recorded as of 10th of January, 2021 (6).

Subsequently, President Cyril Ramaphosa declared a nationwide lockdown on 23 March 2020 to help curb the spread of the COVID-19 in South Africa and encourage health systems to plan for the influx of moderate to severe COVID-19 cases (7, 8). In addition to the national lockdown, other physical distancing steps such as isolation of persons infected with the COVID-19 and quarantining of anyone who might have been exposed or in contact with an infected individual was also encouraged and implemented (9).

Despite the World Health Organization (WHO) advice to national leaders that COVID-19 preparedness efforts should focus on access to “essential medicines” and healthcare services, to prioritise other health needs of the population whilst the Nation is on lockdown (10), some individuals within households and communities in South Africa are deprived of access to essential medicine or health care services, including sexual and reproductive health services, because they feel obligated to uphold the lockdown and prevent transmission of COVD-19 (11).

The strain that the outbreak imposes on health systems will undoubtedly impact the sexual and reproductive health of individuals living in low-and middle-income countries (LMICs) (12, 13) with such consequences as halt the supply of contraceptive products, including condoms, due to restriction imposed as a result of lockdown and physical distancing (14, 15).

Prior to the pandemic, LMICs within sub-Saharan Africa (SSA) and Southern Asia bore the maximum burden of unmet need for modern contraceptives, accounting for 57% of total global unmet needs, of which 39% of these women reside in developing countries (16). Despite the doubling in the number of women consuming modern contraceptive methods from 470 million in 1990 to around 840 million in 2019, an estimated 214 million women in developing countries still had unmet needs for contraceptives currently (16), while South Africa’s overall unmet need for contraception is 18%, with contraceptive prevalence rate (CPR) for married women at 54%, 64% for unmarried women and male condoms use rate of 16% (17).

Guttmacher Institute Authors and other studies estimated that if there were a 10% decline over a year in the use of contraception as a result of inadequate access because of the ongoing pandemic, an additional of over 48 million women would have an unmet need for contraception worldwide, resulting in more than 15 million additional unintended pregnancies (18-20), which may lead to unsafe abortions and higher extra spending in the future on sexual and reproductive health outcomes as a result of coronavirus disease 2019 pandemic (21-23).

However, prior to the COVID-19 pandemic outbreak, multiple factors such as poverty, illiteracy, lack of knowledge and awareness about contraceptives, non-availability of contraceptives and socio□demographic inequalities have been linked to low use of contraceptive (16, 24-26), but the most recent is the limited access to sexual and reproductive healthcare services due to COVID-19 outbreak which made access to their choice of contraception limited as well (27, 28). Limited access to condom use during the COVID-19 outbreak has previously been linked to risky sexual behaviour among nine out of every fifteen adults in Italy (29).

Consequently, with the current evolution of the COVID-19 pandemic, there is a need for concerted actions towards ensuring individuals who need essential access to sexual and reproductive health, including contraception services, are not obstructed (12, 30). Condom use has been recognised as one of the most effective contraceptive methods of preventing unintended pregnancy and sexually transmitted infections (31, 32). Given this dual usefulness, condom services or availability at any point in time should not be obstructed. Thus, there is a need to examine factors associated with condoms access and its sources during the ongoing COVID-19 pandemic in South Africa.

The outcome of this study will be useful to South African health authorities in implementing required interventions that will put into consideration factors contributing to limited access to condom use and preferred source of condom.

## Methods and Materials

### Study design and settings

This study used data from the National Income Dynamics Study-Coronavirus Rapid Mobile Survey (NIDS-CRAM) (33). NIDS-CRAM is a nationally representative survey of the NIDS, which involves a sample of South African from 2017 NIDS wave 5, who were then re□interviewed via telephone interview during the COVID-19 pandemic. This is the first secondary dataset on coronavirus from NIDS during the coronavirus pandemic (34). This survey’s primary investigator is the Southern Africa Labour and Development Research Unit (SALDRU), which is affiliated with the University of Cape Town (UCT). SALDRU is aided by the South Africa Department of Planning, Monitoring and Evaluation (35). The NIDS wave 5 survey employed a stratified, two-stage cluster sample design to interview respondents in all nine South African provinces, and this study maintained the NIDS wave 5 study design and settings for NIDS-CRAM (36).

### Data collection

NIDS-CRAM is a computer-assisted telephone interviewing (CATI) survey, with the first wave conducted during the coronavirus pandemic in South Africa from May to June 2020. Respondents were mainly asked retrospective questions about their circumstances from February to April 2020. The NIDS-CRAM constitutes a sample of 7,074 individuals drawn from the adult sub-sample of the fifth wave of NIDS conducted in 2017. Information such as demographic and economic characteristics, access to condoms and sources of condoms during the COVID-19 pandemic in South Africa were the variables extracted from the NIDS-CRAM wave 1 dataset. The de-identified dataset can be accessed upon request at http:/www.nids.uct.ac.za while the redefined dataset used for this study has been deposited to open science framework (OSF) accessible here https://doi:10.17605/OSF.IO/J4XQR.

### Sampling

After eliminating respondents who failed to answer questions related to access to condoms or source of condoms during the coronavirus pandemic in South Africa, a total of 5,304 respondents were eligible for the study out of 7,074 individuals. The eligible respondents were male and female of reproductive age group between the age of 15 to 49. This is because the reproductive health age group is often defined as those between the ages of 15 and 49. These are the age groups assumed to be sexually active and are majorly in need of sexual and reproductive health services, including condoms (37, 38).

### Statistical Analysis

NIDS-CRAM wave 1 dataset was recoded and analyzed using Stata version 16 software. Descriptive statistics were used to summarize data on demographic characteristics, economic characteristics, access to condoms and preferred sources during the 2019 novel coronavirus pandemic in South Africa. Outcome variables were access to condoms and sources of condoms. Access to condoms was measured by asking the respondents if they have access to condoms (either male or female condoms) during COVID-19 lockdown or not, followed by the source they preferred, while the explanatory variables were demographic and economic characteristics of the respondents. Selected demographic and economic characteristics include; age, population group, gender and province of the respondents, employment status, educational level, and respondents’ wealth quintile. The wealth quintile of the respondents was measured using the nation’s wealth quintile categorization (Upper quintile: R52 078 and above, 4th quintile: R23 156 – R52 077, 3rd quintile: R12 781 – R23 155, 2nd quintile: R7 030 – R12 780, and Lower quintile: R7 029 and below) (39).

Source of condoms has three variable which includes; private sources (respondent getting condoms from the private source like private clinic or hospital), public (respondent getting condoms from the public source like public/government clinic or hospital) and other sources which includes pharmacy shops, road sellers etc.). Dataset was weighted by applying the recommended weight command of “svyset cluster [pw=w1_nc_wgt], strata (stratum)” to avoid over-sampling, and for non-response adjustment, the outputs were summarized as percentages (%) for both explanatory and outcome variables (33). Chi-square was done to check the significant association of the selected demographic and economic variables on access to condoms and sources of condoms, and afterward, binary logistic regression tests were performed to determine adjusted likelihood of the explanatory variables on only access to condoms in the outcome variables; those who had access were coded “1” as “yes” and those who did not have access were coded “0” as “No”. P-value < 0.01, 0.05, 0.001 were considered statistically significant at 95% confidence interval (CI), and explanatory variables with an unadjusted odds ratio (cOR) and adjusted odds ratio (aOR).

### Ethics approval and consent to participate

This study is a secondary analysis of the NIDS-CRAM wave 1 dataset. Ethical approval for NIDS-CRAM was granted by the University of Cape Town (UCT) Commerce Faculty Ethics Committee. In 2017, the NIDS data collectors (Wave 5) conducted a written informed consent process for all participants and only resumed interviews until this procedure had been completed. NIDS-CRAM wave 1 2020 was drawn from the same population sample of wave 5; hence, the participants’ consent was re-validated via telephone interview before proceeding with relevant questions.

## Results

### Percentage distribution of the outcome and explanatory variables

#### Outcome variables

The percentage distribution of the outcome variables presented in figure1 below indicated that 22.57% of the respondents were unable to access condoms during the coronavirus pandemic in South Africa, while 7 out of every 10 respondents preferred public sources, which include public hospital and clinic to get condoms.

**Figure 1:**
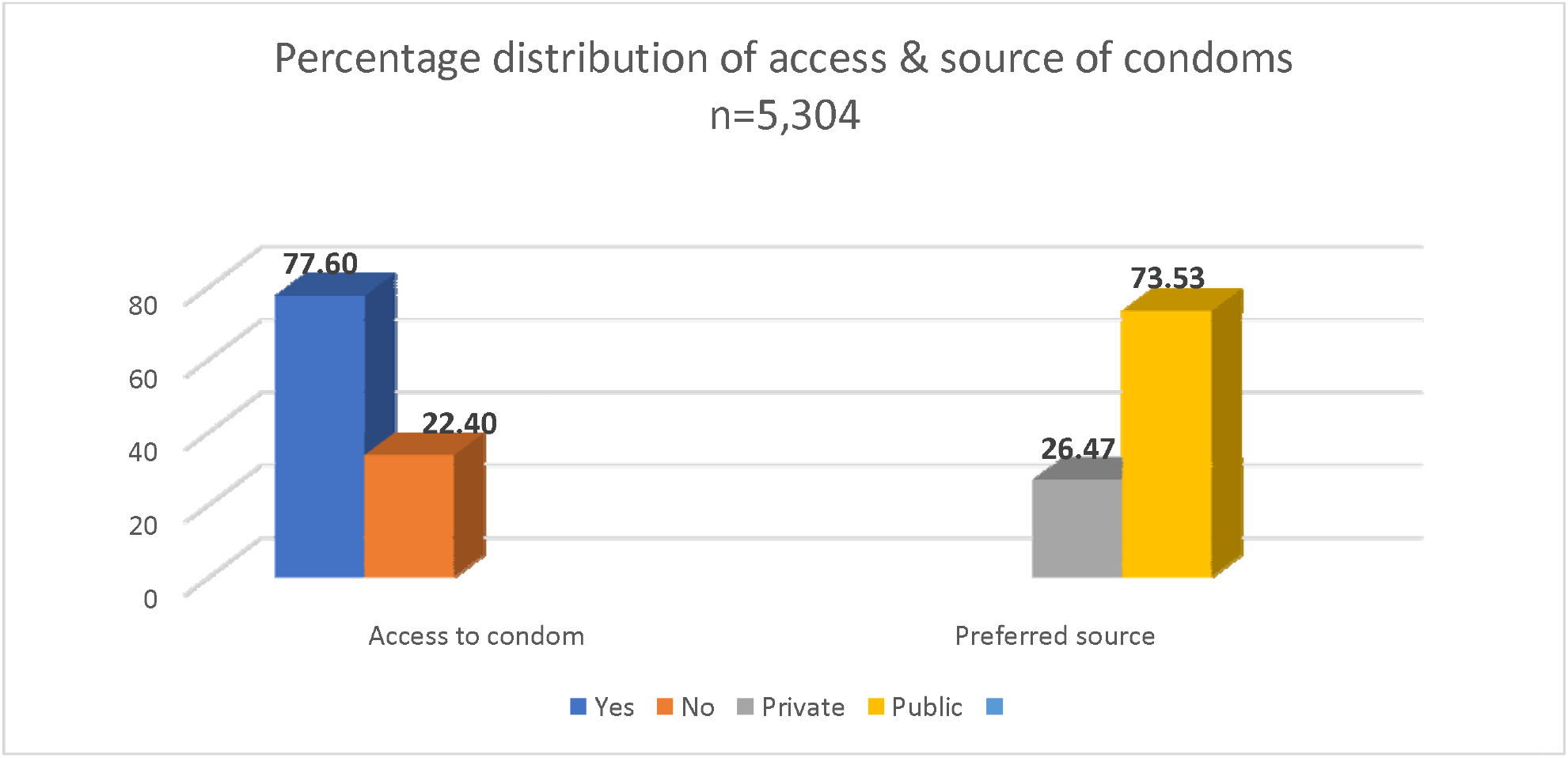
Access and source of condoms.

#### NIDS-CRAM Wave 1, 2020 (Weighted)

Table 1 below showed the percentage distribution of the explanatory variables included in the study.

**Table 1:**
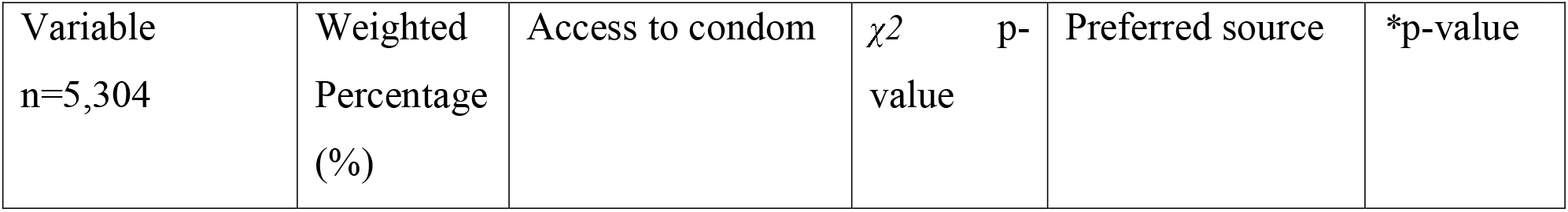
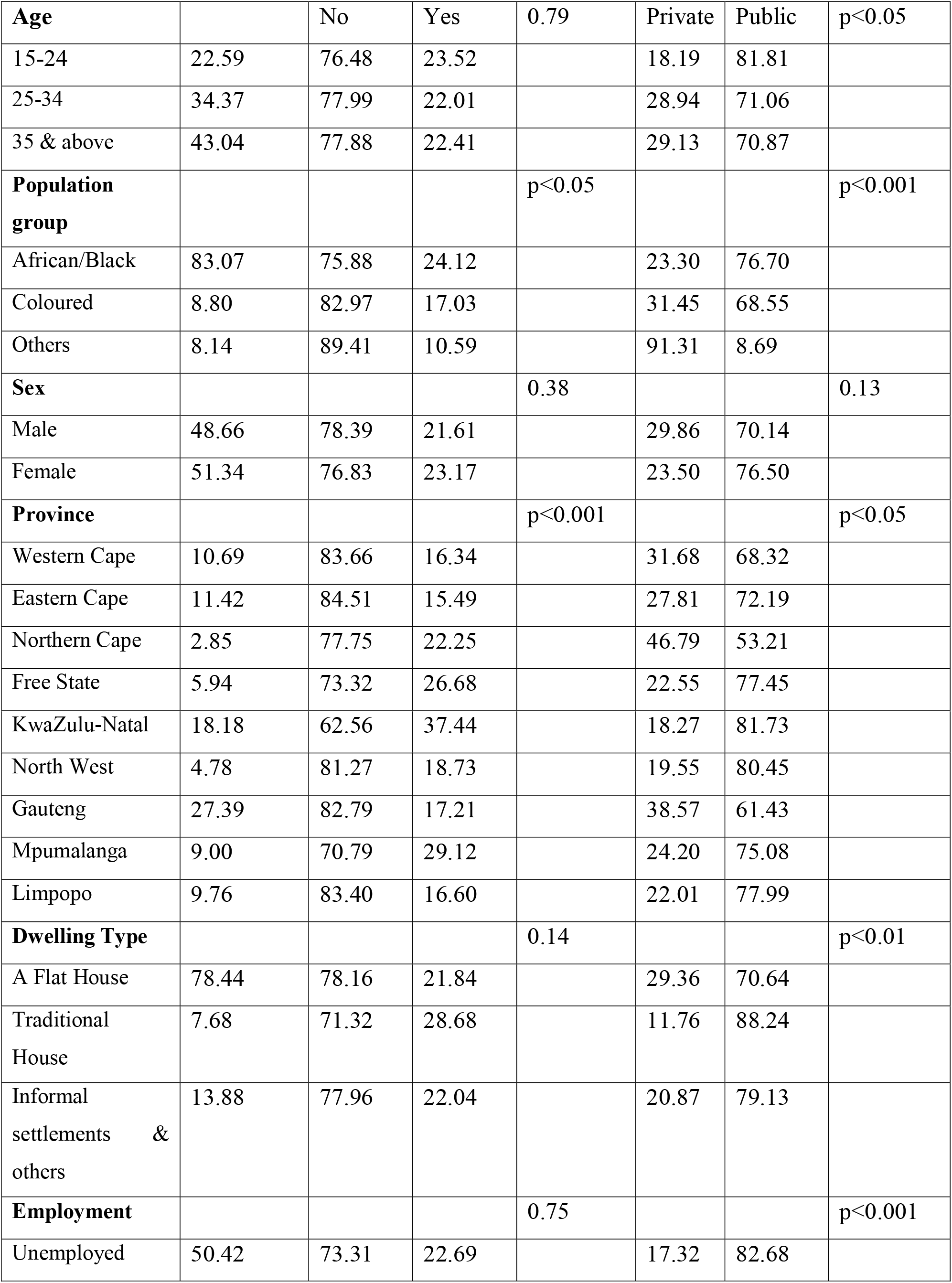
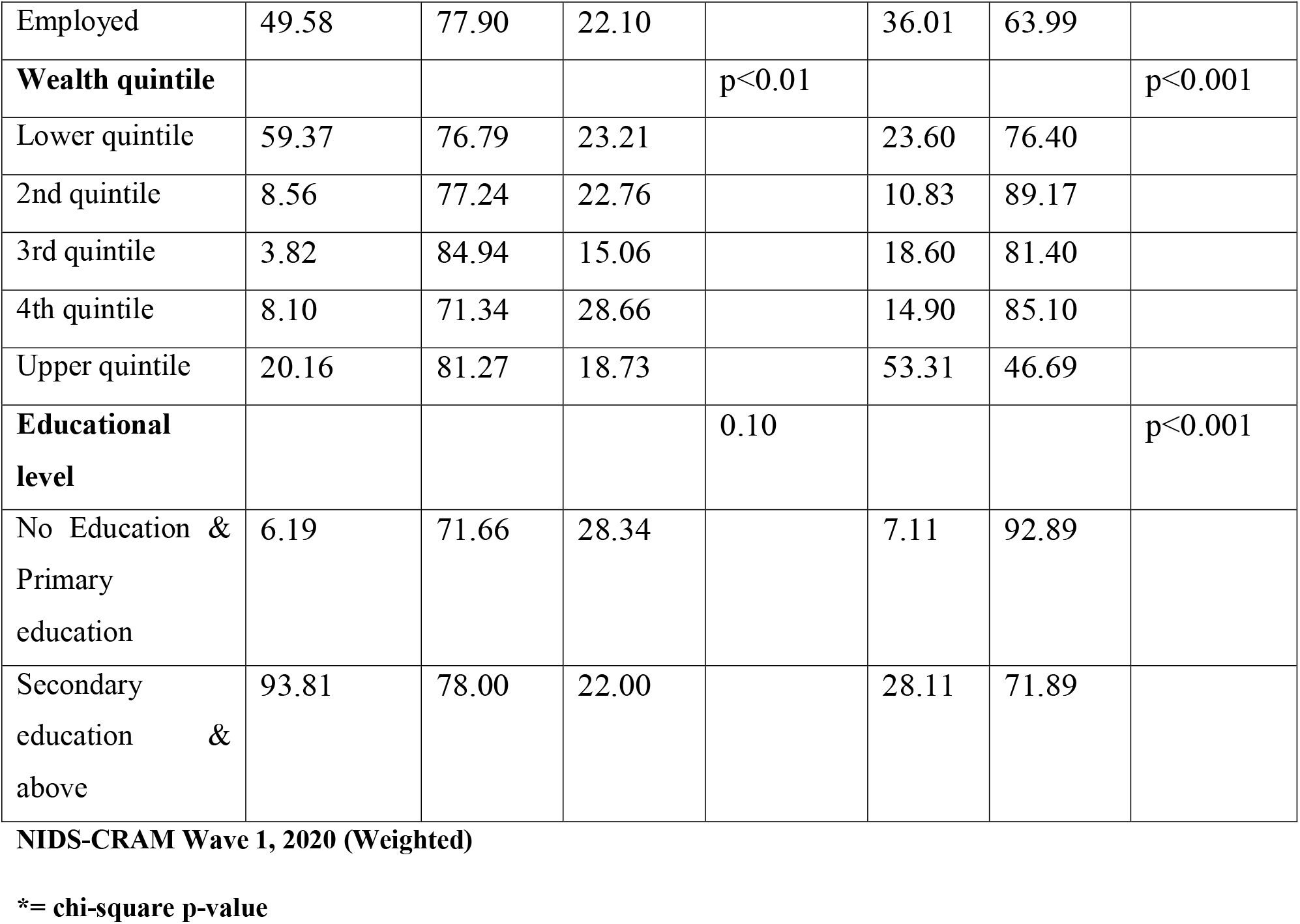
Characteristics distribution of respondents access to condom and preferred source of condom.

The majority (34.37%) of the respondents were between the age group 25-34 years. Almost 8 in 10 (83.07%) of the respondents were Africans or Black, while the lowest population group was among other population groups such as White, Indian and Asian with 8.14%. A little above half (51.34%) of the respondents were female, while males involved in the survey were below average (48.66%). Gauteng had the highest respondents, with 27.39%, followed by KwaZulu-Natal 18.18%, while the lowest was among Northern-Cape (2.85%). Almost 8 in 10 of the respondents dwell in a House or flat residence. 50.42% of the respondents interviewed were employed. 59.37% of the respondents were in the lower quintile. The majority of the respondents had secondary education and above (93.81%).

#### Multivariable Analysis

Table 2 below showed the adjusted multivariate regression results of access condom and preferred source of condom during the COVID-19 pandemic in South Africa.

**Table 2:**
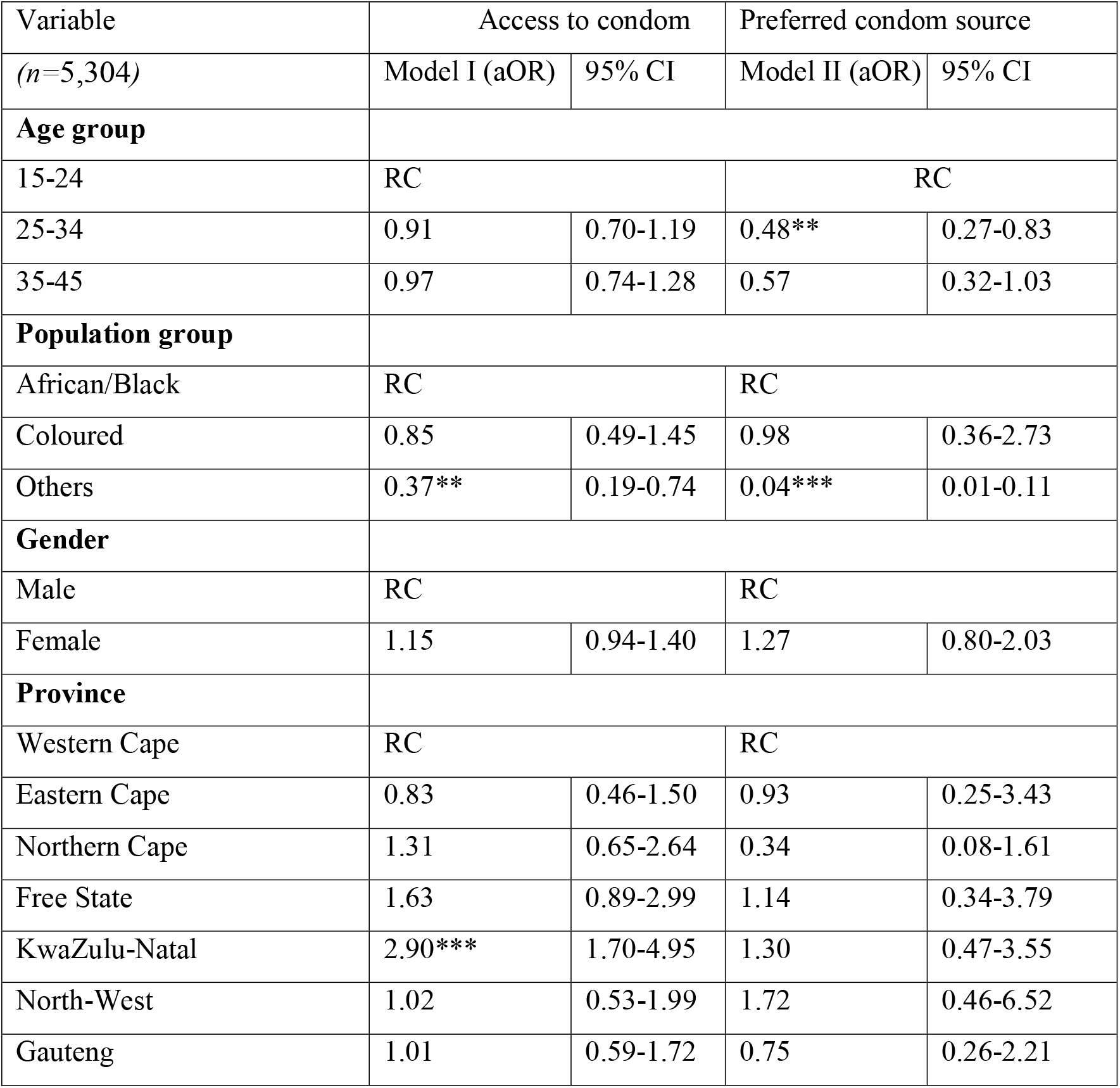
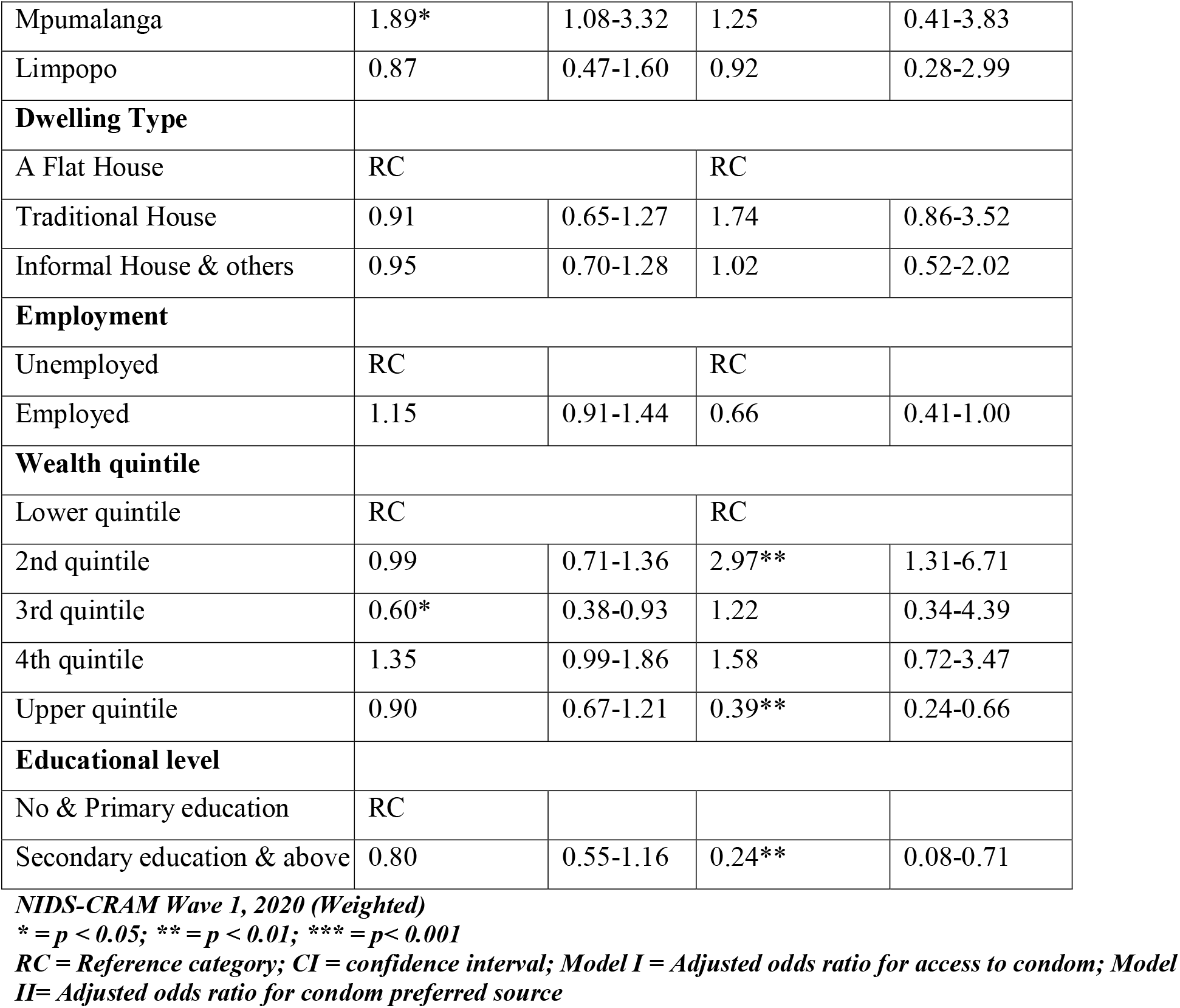
Multivariate logistic regression analysis of factors associated with condom access & preferred source.

| Variable | Access to condom |  | Preferred condom source |  |
| --- | --- | --- | --- | --- |
| (n=5,304) | Model I (aOR) | 95% CI | Model II (aOR) | 95% CI |
| **Age group** |  |  |  |  |
| 15-24 | RC |  | RC |  |
| 25-34 | 0.91 | 0.70-1.19 | 0.48** | 0.27-0.83 |
| 35-45 | 0.97 | 0.74-1.28 | 0.57 | 0.32-1.03 |
| **Population group** |  |  |  |  |
| African/Black | RC |  | RC |  |
| Coloured | 0.85 | 0.49-1.45 | 0.98 | 0.36-2.73 |
| Others | 0.37** | 0.19-0.74 | 0.04*** | 0.01-0.11 |
| **Gender** |  |  |  |  |
| Male | RC |  | RC |  |
| Female | 1.15 | 0.94-1.40 | 1.27 | 0.80-2.03 |
| **Province** |  |  |  |  |
| Western Cape | RC |  | RC |  |
| Eastern Cape | 0.83 | 0.46-1.50 | 0.93 | 0.25-3.43 |
| Northern Cape | 1.31 | 0.65-2.64 | 0.34 | 0.08-1.61 |
| Free State | 1.63 | 0.89-2.99 | 1.14 | 0.34-3.79 |
| KwaZulu-Natal | 2.90*** | 1.70-4.95 | 1.30 | 0.47-3.55 |
| North-West | 1.02 | 0.53-1.99 | 1.72 | 0.46-6.52 |
| Gauteng | 1.01 | 0.59-1.72 | 0.75 | 0.26-2.21 |
| Mpumalanga | 1.89* | 1.08-3.32 | 1.25 | 0.41-3.83 |
| Limpopo | 0.87 | 0.47-1.60 | 0.92 | 0.28-2.99 |
| **Dwelling Type** |  |  |  |  |
| A Flat House | RC |  | RC |  |
| Traditional House | 0.91 | 0.65-1.27 | 1.74 | 0.86-3.52 |
| Informal House & others | 0.95 | 0.70-1.28 | 1.02 | 0.52-2.02 |
| **Employment** |  |  |  |  |
| Unemployed | RC |  | RC |  |
| Employed | 1.15 | 0.91-1.44 | 0.66 | 0.41-1.00 |
| **Wealth quintile** |  |  |  |  |
| Lower quintile | RC |  | RC |  |
| 2nd quintile | 0.99 | 0.71-1.36 | 2.97** | 1.31-6.71 |
| 3rd quintile | 0.60* | 0.38-0.93 | 1.22 | 0.34-4.39 |
| 4th quintile | 1.35 | 0.99-1.86 | 1.58 | 0.72-3.47 |
| Upper quintile | 0.90 | 0.67-1.21 | 0.39** | 0.24-0.66 |
| **Educational level** |  |  |  |  |
| No & Primary education | RC |  |  |  |
| Secondary education & above | 0.80 | 0.55-1.16 | 0.24** | 0.08-0.71 |

The associated factors with access to condoms were other population groups (White, Indian & Asian), KwaZulu-Natal and Mpumalanga provinces and 3rd wealth quintile. Other population groups such as White, Indian & Asian [aOR=0.37; 95% CI=0.19-0.74] and respondents within 3rd wealth quintile [aOR=0.60; 95% CI=0.38-0.93] were less likely to have access to condom compared to respondents who are Black/African and those in the lower wealth quantile while respondents who reside in KwaZulu-Natal [aOR=2.90; 95% CI=1.70-4.95] and Mpumalanga [aOR=1.89; 95% CI=1.08-3.32] provinces were more likely to have access to condom during COVID-19 compared to those residing in Western Cape province.

Factors associated with preferred condom source during the COVID-19 pandemic in South Africa were respondent’s age group, population group, wealth quantile and educational level. Respondents between age 25-34 [aOR=0.48; 95% CI=0.27-0.83], other population groups such as white, Indian and Asian [aOR=0.04; 95% CI=0.01-0.11], respondents within upper quantile [aOR=0.39; 95% CI=0.24-0.66], and those with secondary education and above [aOR=0.24; 95% CI=0.08-0.71] were less likely to preferred public source of condom during COVID-19 pandemic compared to those between the age of 15-24 years, respondents who were Black/African, those within lower wealth quantile, and those with no education/primary education while respondents within 2nd wealth quantile [aOR=2.97; 95% CI=1.31-6.71] were more likely to a preferred public source of condom compared to respondents within lower wealth quintile.

## Discussion

This study examined the factors associated with limited access to condoms and sources of condoms during the COVID-19 pandemic in South Africa using the first national income dynamic study-coronavirus rapid mobile survey (NIDS-CRAM) wave 1 dataset conducted during the pandemic. This study acquired and contributed to the existing literature on how limited access to condoms can increase the unmet need for contraception, which may lead to poor sexual and reproductive health outcomes. The study further expands the scope of the unmet need for contraception by including limited access to condoms during the COVID-19 pandemic and physical distancing in South Africa. In the same vein, the study further contributes to the body of knowledge on preferred source of condoms.

This study is in concordance with the study that concluded that inadequate or limited access to condoms is part of the contributing factors to the unmet need for contraception (40). The result showed that more than two in every ten South Africans experienced limited access to condoms during the pandemic. A study conducted in Indonesia and Kenya on preferred contraception sources showed that most of the population preferred public/government hospitals to obtain contraception prescriptions (41, 42). This is in line with the study result in that more than two-thirds of South Africans preferred public/government hospitals to get condoms. This could be one of the reasons why the respondents were unable to access condoms as most public, or government hospitals were occupied or overwhelmed due to the influx of COVID-19 patients (43, 44) and may also be due to limited transportation as a result of lockdown/ physical distancing (45-47).

As access to contraception continues to be a major contributor to high unmet needs in developing countries (16), this study results were significant to respondents’ population group, provinces, and wealth quintile. South Africans who were White, Asian, and Indian population groups and those in the third wealth quintile were less likely to experience limited access to condoms during the COVID-19 pandemic. This is contrary to the studies conducted in South Africa and Ghana prior to the COVID-19 pandemic that reported that respondents residing in the rural area, those married and female were less likely to have access to condoms (48, 49).

The results on access to condoms further showed that respondents residing in KwaZulu-Natal and Mpumalanga provinces were more likely to have access to condoms during the COVID-19 pandemic. This corroborates with the findings of Ntshiqa, Musekiwa (50), who reported that there was geographical variation in access to condoms in South Africa.

Factors associated with preferred source of condoms include; age of respondents, population group, wealth index and educational level. The result should include that respondents between the age of 25-34 were less likely to prefer public source of condoms. This is in line with a study conducted by Radovich, Dennis (51) that reported that young people prefer private source.

The variation in preferred source of condom reported in this study, in that White, Indian and Asian population group and upper wealth quantile and those with secondary education level and above were less likely to prefer public source of condom while only respondents in 2nd wealth quantile were more likely to prefer public source of condom was similar to a study conducted in Kenya that reported high variation of choice of condom source (42).

Furthermore, this study results showed similarity in most studies, commentaries and editorial opinions that obstruction in sexual and reproductive services in the ongoing COVID-19 pandemic could lead to a high unmet need for contraception (18, 28, 43) and to the best of my knowledge this is the first paper that holistically employed NIDS-CRAM wave 1 dataset to examine factors associated with access to condom and preferred sources of condoms among South Africans during COVID-19 pandemic.

### Strengths and Limitations

The use of secondary datasets has its limitations as some questions of interest to further probe the respondents in terms of retrospective questions were not asked during data collection, and this limited the scope of the study. This study’s strength is the use of aboriginal staff for the telephone interview with the help of computer-assisted telephone interviewing during the COVID-19 pandemic despite the restriction and lockdown.

## Conclusion and Recommendations

Limited access to condoms is the ignored elephant in the room. This study added to the body of literature that there was limited access to condoms during the COVID-19 pandemic and that the preferred source of condoms was very skewed to public source in South Africa.

The study concluded that the demographic and economic characteristics of South Africans influenced their adopted sources of condoms and that limited access to condoms was more experienced among the African/Black population groups, those who reside in Mpumalanga and KwaZulu-Natal provinces and those who were in the third quintile of wealth quintile. Policies, strategies, and interventions such as community distribution of free condoms to avert obstruction access to condom demands of South Africans. This will reduce the unmet need for contraception in South Africa and tackle the unequal family planning use coverage.

## Data Availability

The National Income Dynamics Study-Coronavirus Rapid Mobile Survey (NIDS-CRAM) repressed dataset can be accessed via the NIDS website (http:/www.nids.uct.ac.za). Alternatively, data that includes sensitive information may be obtained via the University of Cape's application process with Dataset (www.datarst.uct.ac.za).

## Abbreviations

COVID-19: Coronavirus 2019
SSA: sub-Saharan Africa
LMICs: Low-and-middle-income countries
NIDS: National Income Dynamic Study
WHO: World Health Organisation
CPR: Contraceptive prevalence rate
NIDS-CRAM: National Income Dynamics Study-Coronavirus Rapid Mobile Survey
SALDRU: Southern Africa Labour and Development Research Unit
CATI: Computer-assisted telephone interviewing
OSF: Open science framework
CI: Confidence interval
COR: Unadjusted odds ratio
AOR: Adjusted odds ratio

## Consent for publication

Not applicable.

## Availability of data and materials

The de-identified dataset can be accessed upon request at http:/www.nids.uct.ac.za while the redefined dataset used for this study has been deposited to open science framework (OSF) accessible here https://doi:10.17605/OSF.IO/J4XQR.

## Author’s contributions

OAB conceptualized the study, wrote the first draft, analyzed & interpreted the results, and overall edited the manuscript and approved the final version

## Acknowledgments

Not applicable

## Authors’ information

OAB: Discipline of Public Health Medicine, College of Health Sciences, University of KwaZulu-Natal, Howard Campus, Durban 4041, South Africa; Obaxlove Consult, Lagos, 100009, Nigeria.

## References

1. Kavanagh MM, Erondu NA, Tomori O, Dzau VJ, Okiro EA, Maleche A, et al. Access to lifesaving medical resources for African countries: COVID-19 testing and response, ethics, and politics. The Lancet. 2020;395(10238):1735–8.

2. Faden YA, Alghilan NA, Alawami SH, Alsulmi ES, Alsum HA, Katib YA, et al. Saudi Society of Maternal-Fetal Medicine guidance on pregnancy and coronavirus disease 2019. Saudi medical journal. 2020;41(8):779–90.

3. Ibarra FP, Mehrad M, Mauro MD, Godoy MFP, Cruz EG, Nilforoushzadeh MA, et al. Impact of the COVID-19 pandemic on the sexual behavior of the population. The vision of the east and the west. International braz j urol. 2020;46:104–12.

4. Caballero AE, Ceriello A, Misra A, Aschner P, McDonnell ME, Hassanein M, et al. COVID-19 in people living with diabetes: An international consensus. Journal of Diabetes and its Complications. 2020;34(9):107671.

5. Nyasulu J, Pandya H. The effects of coronavirus disease 2019 pandemic on the South African health system: A call to maintain essential health services. African Journal of Primary Health Care & Family Medicine. 2020;12(1).

6. (NICD) NIoCD. COVID-19 Surveillance Dashboard 2020 [Available from: https://www.nicd.ac.za.

7. 2020 SAn. Covid-19 News updae 2020 [Available from: https://www.sanews.gov.za/southafrica/president-ramaphosa-announces-nationwide-lockdown.

8. Blumberg L, Jassat W, Mendelson M, Cohen C. The COVID-19 crisis in South Africa: Protecting the vulnerable. South African Medical Journal. 2020.

9. Mukumbang FC, Ambe AN, Adebiyi BO. Unspoken inequality: how COVID-19 has exacerbated existing vulnerabilities of asylum-seekers, refugees, and undocumented migrants in South Africa. International journal for equity in health. 2020;19(1):1–7.

10. Alexander GC, Qato DM. Ensuring access to medications in the US during the COVID-19 pandemic. JAMA. 2020.

11. Joska JA, Andersen L, Rabie S, Marais A, Ndwandwa E-S, Wilson P, et al. COVID-19: Increased Risk to the Mental Health and Safety of Women Living with HIV in South Africa. AIDS and Behavior. 2020:1.

12. Riley T, Sully E, Ahmed Z, Biddlecom A. Estimates of the potential impact of the COVID-19 pandemic on sexual and reproductive health in low-and middle-income countries. Int Perspect Sex Reprod Health. 2020;46:46.

13. Hussein J. COVID-19: What implications for sexual and reproductive health and rights globally?: Taylor & Francis; 2020.

14. Makins A, Arulkumaran S, Contraception F, Committee FP, Sheffield J, Townsend J, et al. The negative impact of COVID□19 on contraception and sexual and reproductive health: Could immediate postpartum LARCs be the solution? International Journal of Gynecology & Obstetrics. 2020.

15. Oyediran KA, Makinde OA, Adelakin O. The Role of Telemedicine in Addressing Access to Sexual and Reproductive Health Services in sub-Saharan Africa during the COVID-19 Pandemic. African Journal of Reproductive Health. 2020;24(2):49–55.

16. (UNFPA) UNPF. Impact of the COVID-19 Pandemic on Family Planning and Ending Gender-based Violence, Female Genital Mutilation and Child Marriage. 2020 [Available from: https://www.unfpa.org/sites/default/files/resource-pdf/COVID-19_impact_brief_for_UNFPA_24_April_2020_1.pdf.

17. Health SADo. South Africa Demographic and Health Survey 2016: Key Indicators Report: Statistics South Africa; 2017.

18. Kumar M, Daly M, De Plecker E, Jamet C, McRae M, Markham A, et al. Now is the time: a call for increased access to contraception and safe abortion care during the COVID-19 pandemic. BMJ Global Health. 2020;5(7):e003175.

19. Ahmed Z, Cross L. Crisis on the horizon: Devastating losses for global reproductive health are possible due to COVID-19, Guttmacher Institute. 2020.

20. Kumar N. COVID 19 era: a beginning of upsurge in unwanted pregnancies, unmet need for contraception and other women related issues. The European Journal of Contraception & Reproductive Health Care. 2020;25(4):323–5.

21. Yazdkhasti M. The novel coronavirus (COVID-19) and unintended pregnancy during the quarantine period. The Pan African Medical Journal. 2020;35(29).

22. Hall KS, Samari G, Garbers S, Casey SE, Diallo DD, Orcutt M, et al. Centring sexual and reproductive health and justice in the global COVID-19 response. The lancet. 2020;395(10231):1175–7.

23. Purdy C. Opinion: How will COVID-19 affect global access to contraceptives–and what can we do about it. Devex Retrieved May. 2020;10:2020.

24. Geldsetzer P, Reinmuth M, Ouma PO, Lautenbach S, Okiro EA, Bärnighausen T, et al. Mapping physical access to healthcare for older adults in sub-Saharan Africa: A cross-sectional analysis with implications for the COVID-19 response. medRxiv. 2020.

25. Okereke M, Ukor NA, Adebisi YA, Ogunkola IO, Favour Iyagbaye E, Adiela Owhor G, et al. Impact of COVID□19 on access to healthcare in low□and middle□income countries: Current evidence and future recommendations. The International Journal of Health Planning and Management. 2020.

26. Desai S, Samari G. COVID□19 and Immigrants’ Access to Sexual and Reproductive Health Services in the United States. Perspectives on Sexual and Reproductive Health. 2020.

27. Mmeje OO, Coleman JS, Chang T. Unintended Consequences of the COVID-19 Pandemic on the Sexual and Reproductive Health of Youth. Journal of Adolescent Health. 2020.

28. Cash R, Patel V. Has COVID-19 subverted global health? The Lancet. 2020;395(10238):1687–8.

29. Balestri R, Magnano M, Rizzoli L, Infusino SD, Urbani F, Rech G. STIs and the COVID-19 pandemic: the lockdown does not stop sexual infections. J Eur Acad Dermatol Venereol. 2020.

30. Tang K, Gaoshan J, Ahonsi B. Sexual and reproductive health (SRH): a key issue in the emergency response to the coronavirus disease (COVID-19) outbreak. Reproductive Health. 2020;17:1–3.

31. Steiner RJ, Liddon N, Swartzendruber AL, Pazol K, Sales JM. Moving the message beyond the methods: Toward integration of unintended pregnancy and sexually transmitted infection/HIV prevention. American journal of preventive medicine. 2018;54(3):440–3.

32. Pazol K, Kramer MR, Hogue CJ. Condoms for dual protection: patterns of use with highly effective contraceptive methods. Public health reports. 2010;125(2):208–17.

33. Kerr A, Ardington C, Burger R. Sample design and weighting in the NIDS-CRAM survey. 2020.

34. Ingle K, Brophy T, Daniels R. National Income Dynamics Study–Coronavirus Rapid Mobile Survey (NIDS-CRAM) panel user manual. Technical Note Version. 2020;1.

35. SALDRU. Development Research Unit. National income dynamics study (NIDS). 2012;2012.

36. Leibbrandt M, Woolard I, de Villiers L. Methodology: Report on NIDS Wave 5. Technical Paper No. 1. Cape Town: Southern Africa Labour and Development Research Unit. 2009.

37. Organization WH. Reproductive Health Indicators–Reproductive Health and Research Guidelines for Their Generation, Interpretation and Analysis for Global Monitoring. Geneva: WHO. 2006.

38. Olagunju OS, Obasanjo BA, Temitope EP, Saliu O, Taiwo I, Musa Z, et al. Does Family Planning Messages Exposure in the Preceding 12 Months Period Predict the Current Use of a Modern Family Planning Method among Women of Reproductive Age in Nigeria? American Journal of Public Health. 2020;8(3):100–4.

39. Stats S. Living conditions of households in South Africa. Avail-18. 2015.

40. Cleland J, Harbison S, Shah IH. Unmet need for contraception: issues and challenges. Studies in family planning. 2014;45(2):105–22.

41. Radhakrishnan U. A dynamic structural model of contraceptive use and employment sector choice for women in Indonesia. US Census Bureau Center for Economic Studies Paper No CES-WP-10-28. 2010:10–01.

42. Jalang’o R, Thuita F, Barasa SO, Njoroge P. Determinants of contraceptive use among postpartum women in a county hospital in rural KENYA. BMC public health. 2017;17(1):604.

43. Ferreira-Filho ES, de Melo NR, Sorpreso ICE, Bahamondes L, Simões RDS, Soares-Júnior JM, et al. Contraception and reproductive planning during the COVID-19 pandemic. Expert Review of Clinical Pharmacology. 2020;13(6):615–22.

44. Hamzehgardeshi Z, Yazdani F, Rezaei M, Kiani Z. COVID-19 as a Threat to Sexual and Reproductive Health. Iranian Journal of Public Health. 2020;49:136–7.

45. Ahmed S, Ajisola M, Azeem K, Bakibinga P, Chen YF, Choudhury NN, et al. Impact of the societal response to COVID-19 on access to healthcare for non-COVID-19 health issues in slum communities of Bangladesh, Kenya, Nigeria and Pakistan: results of pre-COVID and COVID-19 lockdown stakeholder engagements. BMJ Glob Health. 2020;5(8).

46. Brey Z, Mash R, Goliath C, Roman D. Home delivery of medication during Coronavirus disease 2019, Cape Town, South Africa. African Journal of Primary Health Care & Family Medicine. 2020;12(1):4.

47. Olagunju OS, Bolarinwa OA, Babalola T. Social distancing, lockdown obligatory, and response satisfaction during Covid-19 pandemic: perception of Nigerian social media users. 2020.

48. Marrone G, Abdul-Rahman L, De Coninck Z, Johansson A. Predictors of contraceptive use among female adolescents in Ghana. African journal of reproductive health. 2014;18(1):102–9.

49. Chimbindi NZ, McGrath N, Herbst K, San Tint K, Newell M-L. Socio-demographic determinants of condom use among sexually active young adults in rural KwaZulu-Natal, South Africa. The open AIDS journal. 2010;4:88.

50. Ntshiqa T, Musekiwa A, Mlotshwa M, Mangold K, Reddy C, Williams S. Predictors of male condom use among sexually active heterosexual young women in South Africa, 2012. BMC public health. 2018;18(1):1–14.

51. Radovich E, Dennis ML, Wong KL, Ali M, Lynch CA, Cleland J, et al. Who meets the contraceptive needs of young women in sub-Saharan Africa? Journal of Adolescent Health. 2018;62(3):273–80.

